# The Association between Emotional Intelligence and Depression among Medical Students in Suez Canal University

**DOI:** 10.1101/2022.08.31.22279446

**Authors:** Nada M. El Garhy, Ghada Hegazy, Merna Shata, Mohamed Ibrahim, Hadeel Abdel Kariem, Alaa Eleleedy, Soliman Magdy, Kareem Mohamed

## Abstract

**Background:** Emotional intelligence is defined as the capacity to be aware of, control, and express one’s emotions, and to handle interpersonal relationships judiciously and empathetically while depression is the loss of interest in daily activities that may be associated with altered dietary and sleep patterns, lowered self-esteem, feelings of guilt and worthlessness and recurrent thoughts of suicide.

Previous studies that assess depression among medical students show variable results, which makes it difficult to conclude whether depression rates are higher exclusively in medical students. Moreover, previous studies have shown that emotion management leads to less stress and may allow for adaptation by allowing subjects to induce a more positive mood and may even prevent development of a mal-adaptive emotional state.

**Aim:** To improve mental health of medical students by identifying and modifying risk factors that may contribute to its deterioration

**Methods:** Our sample size is 149 students which were picked by stratified random sampling, where stratification was done on gender and study year. Students’ names of all six years were collected and then split according to study year, each study year was further splitt into two groups of males and females, which were selected from, based on a computerized random sampling. Two questionnaires were used; “Trait Emotional Intelligence Questionnaire” (TEIQue) developed by K. V. Petrides, PhD, for emotional intelligence and Beck’s Inventory for depression measurement.

**Results:** Our sample is composed of 82 (response rate is 55%) students. Depression prevalence in our sample is 39%. And it significantly correlated negatively with emotional intelligence, where the most significant factor is well-being.

**Conclusion:** Stressful lifestyle of medical students and lack of extracurricular activities and skill training programs lead to lowered emotional intelligence levels and depression. We recommend that their should be psychiatric counselling available for medical students, to lower depression prevalence among them, and introduce to curriculum life skill training programs to increase levels of emotional intelligence. We also recommend further studies about risks and causes of depression in Suez Canal University medical students to lower its prevalence, contributing to a better quality of life for the future physicians and thus of the patients.

---

Emotional Intelligence(EI) can be defined as the ability to grasp, appreciate and discerningly manage one’s own and other people’s emotions, and then use emotional information to guide thinking and behavior^12^. In 1997 Mayor and Salovey offered the 4 - branch model of EI, stating that EI involves the ability to: perceive, appraise and express emotions; use emotions to facilitate thinking; understand and analyze one’s own and others’ emotions; and regulate emotions reflectively to promote emotional and intellectual growth^3^. The Intelligent Quotient (IQ) was hitherto taken as the standard of excellence in life, but Goleman clarified why EI may matter than IQ.^4^ Moreover, in 1999, Mayer J. D., Caruso D. R., & Salovey P. showed that EI met the criteria of the standard intelligence, highlighting the importance of EI and its effect on mental health^5^. More recently in 2014, a study done on 152 patients with focal brain injuries to obtain measures of emotional intelligence, general intelligence and personality using the Mayer, Salovey, Caruso Emotional Intelligence Test (MSCEIT), the Wechsler Adult Intelligence Scale and the Neuroticism-Extroversion-Openness Inventory, respectively revealed that low scores for measures of general intelligence and personality were related to low scores of emotional intelligence,^6^ proving the importance and the great influence of EI on mental health.

The American Psychological Association defines depression as the loss of interest in daily activities that may be associated with altered dietary and sleep patterns, lowered self-esteem, feelings of guilty and worthlessness and recurrent thoughts of suicide.^7^ Previous studies that assess depression among medical students show variable results, which makes it difficult to conclude whether depression rates are higher exclusively in medical students. A review on previous studies that compared medical to non-medical students in terms of depression showed that non-medical students had a higher prevelance of depression than medical students.^8^ Another study showed that 3% of medical students had suspected depressive disorder, and only 0.5% had depressive disorder.^9^

On the other hand, in North America, the rate of depression in medical students rangedfrom 6.0 to 66.5%^10^. In China the rates of depression and suicidal ideation in medical students were 13.5% and 7.5%, respectively;^11^ while in a private medical school in India, depression prevelance among medical students was 64%.^12^

Additionally, a study that was done in Serbia suggests that medical students are a “at-risk” population to depression.^13^ 92

The public appeal of EI and the development of EI training programs began long before a stronger relationship was established between EI and mental health. ^14^ However, with the development of valid modes of EI assessment, such as Schutte’s self-report questionnaire, the obvious question became the effect of EI on mental health, as measured by stress, depression, and suicide ideation. ^15^ Previous studies have shown that emotion management leads to less stress and may allow for adaptation by allowing subjects to induce a more positive mood^16^and may even prevent development of a maladaptive emotional state.^17^ In addition, a high EI has been connected to better interpersonal skills and allow for greater social support, which may have a protective effect from somatic disease.^18^ On the other hand, a lower EI that manifests as a lack of emotion recognition and/or management is a key sign of some personality disorders and may be linked to impulse control problems. ^15^ Individuals with a high EI may have better emotion management and greater impulse control than their counterparts with a lower EI, but a 2003 study found that such individuals may be more susceptible to mood induction procedures, even negative inductions, leading to stress mismanagement and exacerbation under demanding circumstances. ^19^

Still, more critical work is needed to examine the potential relationship between EI and mental health and whether EI is a distinctive predictor of emotion and stress management, depression, and suicidal ideation. This study seeks to add to the small body of work done on the subject.

## SUBJECTS AND METHOD

### STUDY DESIGN

Observational Cross-sectional study

### STUDY SETTING

Faculty of Medicine, Suez Canal University

### STUDY POPULATION

Medical Students

#### Inclusion /Exclusion Criteria 128

All undergraduate medical students are eligible for the study

### SAMPLING

#### Sample size

149

#### Sampling technique

Our sample is Stratified random sample, where stratification was done on gender and study year. Students’ names of all six years were collected and then split according to study year, each study year was further split into two groups of males and females, which will be selected from, based on a computerized random sampling.

## METHODS

### Exposure variable(s)

Emotional Intelligence: measured using “Trait Emotional Intelligence Questionnaire” (TEIQue) developed by K. V. Petrides, PhD. It is a scale-based questionnaire assessing 15 facets of EI. Average of all subscales is summed into a percentage (quantative variable).

### Outcome variable(s)

Depression: measured using Beck’s Inventory, which is a scale-based inventory. The final score (quantitative variable) is interpreted by the scoring index into degree of depression (qualitative variable)

### Data Collection Tools

- Pen and Pencil self-report questionnaire.
- Online questionnaire.

156

### Procedures

Students were contacted mainly via Social Media (Facebook), and if the student was 158 not reachable then he was contacted in person.

## RESULTS

Our sample is composed of 82 students, 52 female and 30 male students, taken proportionally from each study year using stratified random sampling technique.Depression prevalence in our sample is 39% (figure 1). Prevalence of depression among male and female students is 40% and 38.5%, respectively (Table 1). Beck’s Depression Inventory scoring system divides depression levels into four categories: Minimal depression, mild depression, moderate depression and severe depression. The rate of students in each category is presented in table 2. Trait emotional intelligence questionnaire intereprets five personality factors, which are: Global trait EI, well-being, sociability, emotionality and self control. These factors are further subdivided into 15 facets. When correlating EI factors and depression, results showed significant negative correlation (table 3, Figure 2). However when correlating all 15 facets with depression, all of them showd significant negative correlation except emotion regulation, emotion expression, empathy, relationship and adaptability (Table 4)

**Table 1:**
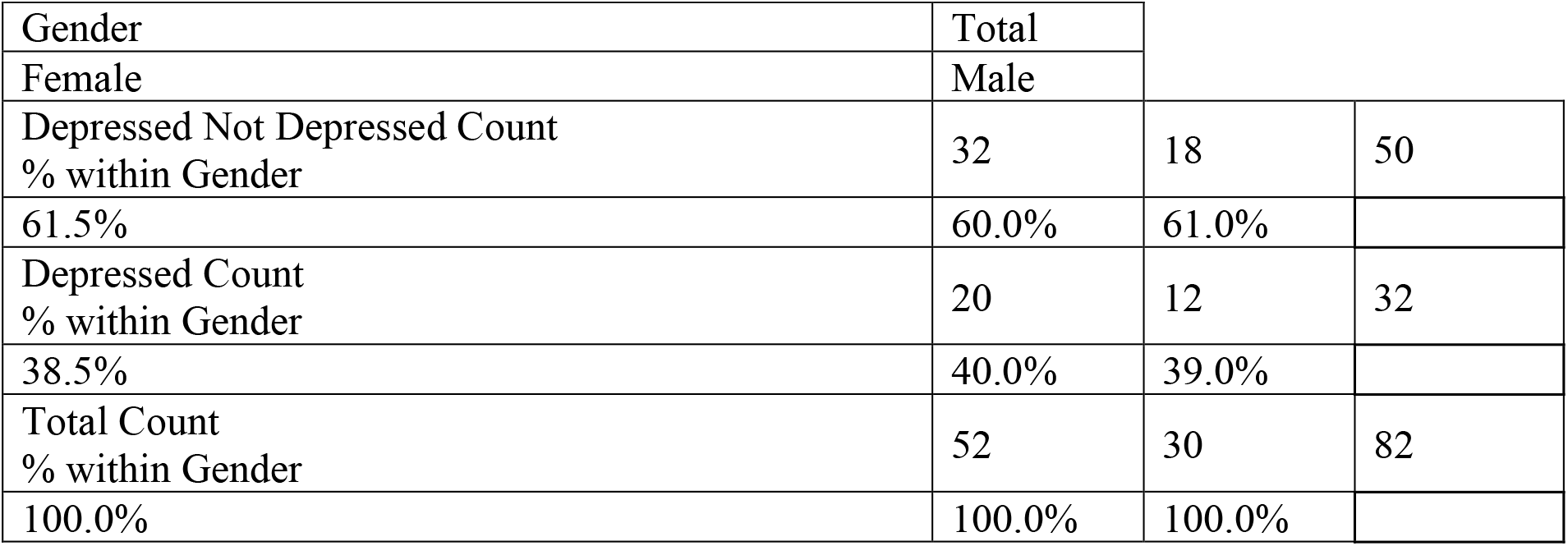
Depressed students according to gender.

**Table 2:**
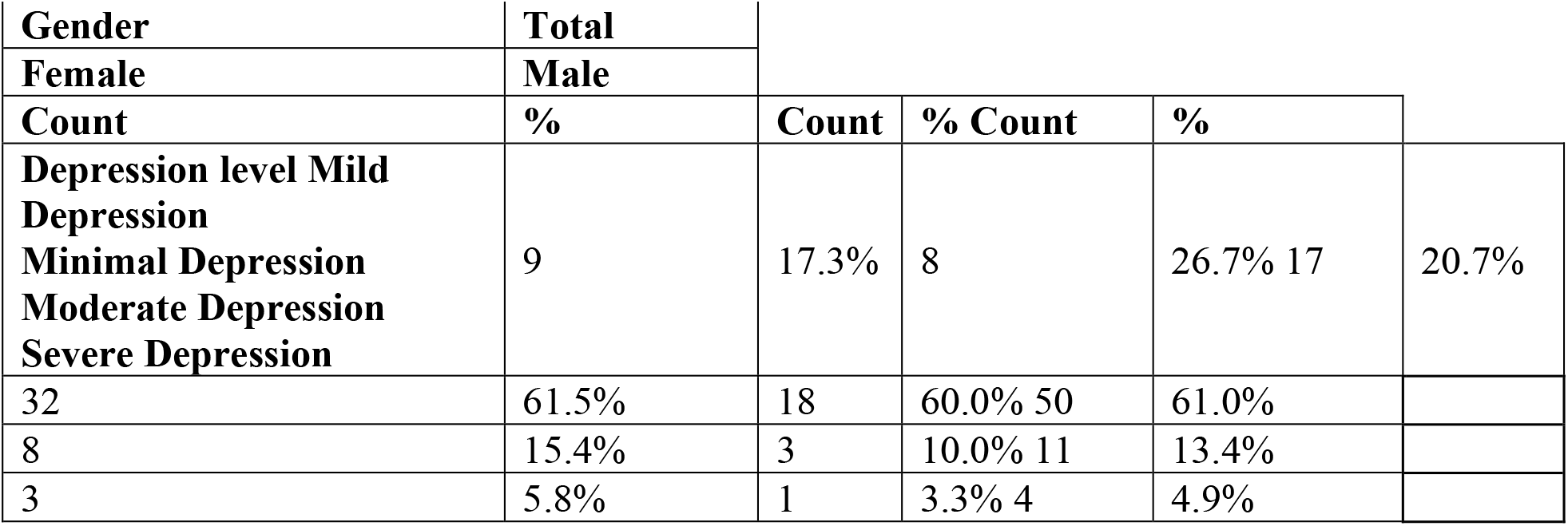
Depression severity according to gender.

**Table 3:** Correlation between Emotional intelligence and depression.

| Subgroup | Depression |
| --- | --- |

| Pearson Correlation | Sig. (2-tailed) |  |
| --- | --- | --- |
| global trait EI | -.474** | <0.001 |
| well being | -.593** | <0.001 |
| self-control | -.332** | 0.002 |
| Emotionality | -.292** | 0.008 |
| Sociability | -.299** | 0.006 |
| ** Correlation significance is less than 0.01 |  |  |

**Table 4:**
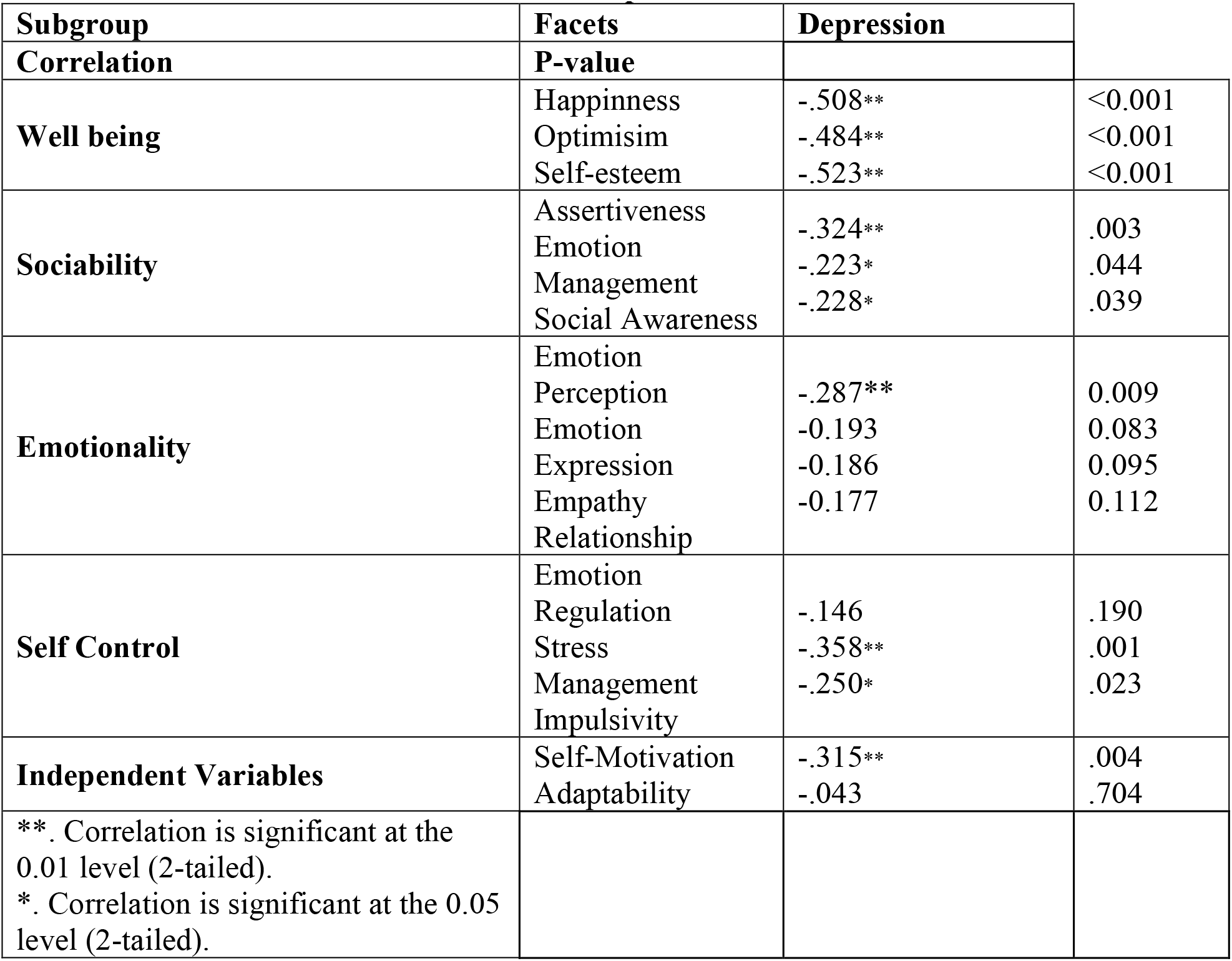
Correlation between EI facets and Depression.

## DISCUSSION

There are two models of Emotional Intelligence that could be conceptualized: the ability model and the trait model. Ability EI, as defined by Mayer &Salovey in 1997, is “ the ability to perceive and express emotion, access and generate feelings when they facilitate thought, understand emotional knowledge, and regulate emotionsto promote intellectual growth, “^3^ whereas trait EI is defined as a constellation of self - perceptions located at the lower levels of personality hierarchies.^20^ A comprehensive meta-analysis of the relationship between emotional intelligence and health done in 2009 on 19,815 participants revealed that EI was more strongly associated with mental health when measured as a trait than when it was measured as an ability.^21^

Based on this and other studies we operationalized the EI as the trait model using the self-reported trait emotional intelligence questionnaire (TEIQue). The TEIQue consists of 15 facets, organized in 5 factors: global trait EI, well-being, self-control, emotionality, and sociability, all of which showed negative correlation with depression.

Several studies have shown that emotional intelligence is ofsignificant relevance to all components of health, including physical health, if to a lesser degree.^22^Stigma towardsdepression and suicide among medical students was explored in various studies, allowing for the unresolved stress in addition to their refusal to seek help.^23^ Our results are similar to several studies done to assess the relationship between EI and depression (but not in medical students), including a study done on high school students in the US ^24^ and another on adolescents in Spain^25^that used Trait Meta mood

Scale (TMMS) instead of the TEIQue, which revealed negative association. A similar study done on diagnosed patients of depression in Lahore, Pakistan showed that low emotional intelligence is a risk factor for depression.^26^ When correlating TEIQue factors to depression, the most significant factor was well-being (p value <0.001). All facets of well-being (happiness, optimism, and self-esteem) showed significant negative corelation wih depression. Previous studies have found that all of these facets are directly related to depression. In the case of omptimistic individuals, they tend to handle stressful situations in a positive manner; they know how to cope and adapt to changing circumstances, unlike pessimistic individuals, leaving them exposed to psychopathy and depression.^27^ In previous studies, students reported the greatest depression levels related to experiencing interpersonal concerns (troubled friends or family members, death of a friend or family member, and relationship difficulties) and mental health concerns (depression, anxiety, seasonal affective disorder and stress)^28^; these affect the facets of happiness and self-esteem negatively. Also, academic achievements may have an 251 important role in building self-esteem. A study comparing EI trends of medical residents and the general population found that the residents scored higher in self-control.^29^Self-control is the regulation of external pressure, emotion, stress, and impulses ^19^ and was significantly correlated to depression in our study (p value < 0.05). Its association with the medical profession can be explained by a combination of self-selection and training that teaches students to regulate emotions and impulsive behavior.^29^Upon closer examination, stress management (p value = 0.001) and impulsivity (p value < 0.05), two of the three facets included under self-control, were statistically significant, with stress management being the most significant to depression. This is in accordance with previous studies and can be explained with Beck’s cognitive theory of depression.^14,30,31^

The majority of medical students are subjected to great external stress in the form of a heavy course load in a limited time frame, a stressor that calls upon essential skills in the management of stress and impulsive behavior that may cause them to abandon their studies for more pleasurable pursuits. Beck’s cognitive theory of depression hypothesizes that depression occurs secondary to negative thought processes centered around a negative cognitive triad involving the self, the world, and the future.This triad yields three dysfunctional self-schemas: one becomes convinced one is defective (self), with all experiences resulting in failure (world), and their future rendered hopeless (future). When applied to all thinking, these self-schemas create three defective thought processes: selective abstraction, which is the perception of negative outcomes only while ignoring any positive results; polar reasoning, the “all-or-nothing” mentality that renders anything less than perfect as a failure in one’s eyes; and over-generalization, the rationalization that a previous failure will result in future failures forever. ^32^ We can thus speculate that individuals with low stress management scores may be unable to cope with the demands on medical school, leading to failure that may stimulate negative cognition and result in depression. On the other hand, low impulse control may adversely affect academic performance, leading to failure that may also stimulate the defective thought processes preceding – and causing – depression.

Another important factor of emotional intelligence is sociability, the tendency to seek social interaction ^33^. It can be divided into two categories: (A) behaviors that bring people together, such as social bonding and (B) behaviors that separate people, such as aggressive behavior. On the surface, both behaviors appear as opposite ends of the social spectrum, and one might be preferred to the other, but from an evolutionary standpoint, each extreme is essential for survival; affiliative behaviors are essential for mating and herd formation, which increases the sense of security to relief stress and anxiety. On the other hand, aggressive behaviors are also essential to maintain that herd and ensure its hierarchy as well as protect the group; thus, relieving stress and anxiety. So, theoretically, the two extremes of the sociability spectrum should ultimately reduce the risk for depression. This particular personality trait may affect medical students management techniques and response toward stress, leading them to depression, affecting their performance professionally ^34–36^

Recent studies have shown that personality traits concerning sociability i.e. extraversion, self-esteem and neuroticism, to be second in order as predictors for student academic expectations and performance after conscientiousness ^37^, which support our findings concerning sociability being the third biggest predictor for the risk depression after state of well-being and self-control (P-value 0.006).

Another 12-year longitudinal study on medical students in the UK, with sample size of 1,668 ^38^ proposed that personality factors mediate physicians’ approaches to work and learning, leading to stress. It also pinpointed that certain traits affect stress susceptibility more than others i.e. high levels of neuroticism and self-awareness, low levels of extraversion, and low levels of conscientiousness. This was supported also by another study one in four-year medical schools in Norway^39^ The emotionality factor showed significant negative correlation with depression; however, of its facets - emotion perception and expression, relationship and empathy-only emotional perception showed significant negative relationship.

Emotional perception is defined as recognition of one’s – and others’ - emotions.^40^ Emotions are initially cognized through visual, auditory, olfactory, or somatic stimuli, leading to a physiological arousal and translated into an emotion, which may then be expressed or suppressed.^41^ According to this, depression is a form of expression of one’s emotions, which proposes whether low scores in emotional perception are the reason students are depressed or is the other way round, i.e. depression eventually leads to a lack of misunderstanding one’s and others’ emotions. Beck’s mood-congruity theory proposed that when one is depressed, one perceives others’ emotions negatively, and when one is in a good mood, emotions are perceived positively. ^42^

When one perceives negative emotions, the feeling of loss of power and willingness in coping with such circumstances in a positive manner is called depression ^43^ In addition, decreased emotional perception may lead to misunderstanding self and other emotions, thereby corrupting healthy social interaction, which may precipitate depression.^44^ On the contrary, decreased emotional expression from others -which is perceived by oneself-could lead to emotional insensitivity, making sociability and peer interactions more difficult to correctly construct, leading ultimately to depression.^45^

Students perceive negative emotions on a daily basis in different ways, whether from their peers, academic performances, or even patients. The inability to regulate such emotions in a positive manner precipitates depression. Another reason is that clinical students are prone to long periods of negative emotion perceived from patients, all while suppressing their own emotions, leading to emotional insensitivity, which leads to lack of emotional perception. Such lack of recognition and expression eventually yields a corrupted peer-peer relationship. In addition, the high prevalence of depression among medical students aggravates the feeling of negative emotions in the general environment. The current literature shows increased interest in proving the increased psychological distress among medical students, as emphasized in this systematic review of 40 studies done on U.S. and Canadian medical students^46^, but still the information available is insufficient to draw a concrete conclusion on the exact causes.

Another review of 17 studies sought to link depression to decreased academic and professional performance.^37^. The literature has been used to describe the relationships between personal traits, stress, depression, and academic performance; however, it has not covered the relationship between emotional intelligence and depression, which has been specifically presented in our study.

## LIMITATIONS

Several limitations of our study must be mentioned. This study was conducted with self-report measures, so it is likely that social desirability may have influenced the responses. The cross-sectional design of our study should be also considered in assumption causality and a follow-up cohort study would be valuable to address this limitation. Finally, the low response rate (51%) made it difficult to make our sample size large enough for further comparison between male and female or study year.

## CONCLUSION AND RECOMMENDATION

Stressful lifestyle of medical students and lack of extracurricular activities and skill training programs lead to lowered emotional intelligence levels and depression. In addition, emotional intelligence levels were negatively correlated with depression. We recommend that there should be a psychiatric counseling available for medical students, to lower depression prevalence among them, and introduce to curriculum life skill training programs to increase levels of emotional intelligence. We also recommend further studies about risks and causes of depression in Suez Canal University medical students to lower its prevalence, contributing to a better quality of life for the future physicians and thus of the patients.

## Data Availability

All data produced in the present study are available upon reasonable request to the authors

## ACKNOWLEDGEMENT

We would like to express my sincere appreciation and deepest gratitude for the cooperation and generous help of the participants. We would also like to thank our mentors (Dr. Ahmed Fouad and Dr. Yosra Saeed; Lecturers of Community department) for their continuous support and valuable tips.

## ETHICAL CONSIDERATION

Human subject’s names will be kept on a password protected database and will be linked only with a study identification number for this research. There are no patient identifiers. All data will be entered into a computer that is password protected. This study does not present any direct benefit to the participants. However the study does provide an opportunity to gain a better understanding of how Emotional Intelligence and depression are related in medical students, and whether gender plays a role in that relationship or not.

